# Evolving epidemiology of novel coronavirus diseases 2019 and possible interruption of local transmission outside Hubei Province in China: a descriptive and modeling study

**DOI:** 10.1101/2020.02.21.20026328

**Authors:** Juanjuan Zhang, Maria Litvinova, Wei Wang, Yan Wang, Xiaowei Deng, Xinghui Chen, Mei Li, Wen Zheng, Lan Yi, Xinhua Chen, Qianhui Wu, Yuxia Liang, Xiling Wang, Juan Yang, Kaiyuan Sun, Ira M. Longini, M. Elizabeth Halloran, Peng Wu, Benjamin J. Cowling, Stefano Merler, Cecile Viboud, Alessandro Vespignani, Marco Ajelli, Hongjie Yu

## Abstract

**Background:** The COVID-19 epidemic originated in Wuhan City of Hubei Province in December 2019 and has spread throughout China. Understanding the fast evolving epidemiology and transmission dynamics of the outbreak beyond Hubei would provide timely information to guide intervention policy.

**Methods:** We collected individual information on 8,579 laboratory-confirmed cases from official publically sources reported outside Hubei in mainland China, as of February 17, 2020. We estimated the temporal variation of the demographic characteristics of cases and key time-to-event intervals. We used a Bayesian approach to estimate the dynamics of the net reproduction number (*Rt*) at the provincial level.

**Results:** The median age of the cases was 44 years, with an increasing of cases in younger age groups and the elderly as the epidemic progressed. The delay from symptom onset to hospital admission decreased from 4.4 days (95%CI: 0.0-14.0) until January 27 to 2.6 days (0.0-9.0) from January 28 to February 17. The mean incubation period was estimated at 5.2 days (1.8-12.4) and the mean serial interval at 5.1 days (1.3-11.6). The epidemic dynamics in provinces outside Hubei was highly variable, but consistently included a mix of case importations and local transmission. We estimate that the epidemic was self-sustained for less than three weeks with *Rt* reaching peaks between 1.40 (1.04-1.85) in Shenzhen City of Guangdong Province and 2.17 (1.69-2.76) in Shandong Province. In all the analyzed locations (n=10) *Rt* was estimated to be below the epidemic threshold since the end of January.

**Conclusion:** Our findings suggest that the strict containment measures and movement restrictions in place may contribute to the interruption of local COVID-19 transmission outside Hubei Province. The shorter serial interval estimated here implies that transmissibility is not as high as initial estimates suggested.

## Introduction

Since December 2019, an increasing number of atypical pneumonia cases caused by severe acute respiratory syndrome coronavirus 2 (SARS-CoV-2) have been reported in Wuhan City of Hubei Province, China^1^. As of February 17, 2020, a total of 72,436 cases of novel coronavirus diseases 2019 (COVID-19) with 1,868 deaths have been reported in mainland China^2^. The outbreak has now spread to twenty-five countries beyond China^3^. The World Health Organization (WHO) declared the COVID-19 outbreak as a Public Health Emergency of International Concern on January 30, 2020^4^.

An early report on epidemiology of the COVID-19 outbreak included analysis of the first 425 confirmed cases detected in Wuhan by January 22, 2020^5^. Since then, the temporal epidemic curve and spatial dissemination of COVID-19 changed quickly, rapidly growing to 72,436 cases by February 17 with 17% of cases reported outside of Hubei. In these other provinces, the COVID-19 epidemic is characterized by a mix of local transmission and importation of cases from Hubei^6^. A recent report based on the 44,672 confirmed cases detected in mainland China by February 11 gave a description of the characteristics of COVID-19 cases for China overall and for Hubei Province^6^. However, until now, there has been little information to understand the epidemiological features and transmission dynamics of COVID-19 outbreak beyond Hubei. This information would be crucial to inform intervention policy in real-time not only for China but also for other countries that are receiving cases from China.

In the present study, we aim to describe the epidemiological characteristics of COVID-19 outbreak after 50 days of detection in the provinces outside Hubei. We also estimate changes in key time-to-event distributions and reproduction numbers to assess whether the strict control measures have been able to slow transmission.

## Methods

### Case definitions and surveillance

Since the outbreak of atypical pneumonia cases was detected in Wuhan at the end of December 2019, the Chinese Center for Disease Control and Prevention (China CDC) has launched a new surveillance system, first in Wuhan, then extended to the entire country, to record information on COVID-19 cases. Case definitions for suspected cases and laboratory-confirmed cases, and the description of the surveillance system have been published^1^ and reported elsewhere^5^. Details are summarized in Appendix (page 2).

In the first version of “Guideline on diagnosis and treatment of novel coronavirus infected pneumonia (NCIP)” issued by China CDC on January 15, 2020, a suspected NCIP case was defined as pneumonia that fulfilled clinical criteria (fever; radiographic findings of pneumonia; or normal or reduced white blood cell count, or reduced lymphocyte count at early onset of symptoms) and had an epidemiologic link to the Huanan Seafood Wholesale Market in Wuhan or travel to Wuhan within 14 days before symptom onset. The subsequent two versions of case definitions issued on January 18 and 22, separately, removed one of the clinical criteria (i.e. no reduction in symptoms after antimicrobial treatment for 3 days following standard clinical guidelines) to accelerate identification of cases and revised the epidemiological link (e.g. a travel history to Wuhan or direct contact with patients from Wuhan who had fever or respiratory symptoms within 14 days of symptom onset, or to be a potential case in a cluster). In the fourth version issued on January 27, the clinical criteria was loosened to meet any two of the three prior clinical criteria (i.e. fever; radiographic findings of pneumonia; normal or reduced white blood cell count, or reduced lymphocyte count at early stage of illness) while the epidemiological link added one more criteria (e.g. epidemiological link with confirmed COVID-19 case). In the fifth version issued on February 4, clinically-diagnosed cases were defined as suspected cases with radiographic findings of pneumonia, to be used exclusively in Hubei^7^.

### Data collection

Daily aggregated data on the number of cumulative cases in mainland China were extracted from the official websites of national, provincial, and municipal Health Commissions (see Tab. S1 in Appendix).

Individual records on laboratory-confirmed COVID-19 cases were collected from two official publicly available sources, including: (i) websites of national, provincial, and municipal Health Commission; (ii) websites of national and local government affiliated medias. Individual information was extracted and entered into a structured database comprising demography, exposure and travel history, timeline from exposure, symptom onset, hospital admission, and date of official announcement (reporting date). Each individual record was extracted and entered by three coauthors and was cross-checked to ensure data accuracy. Conflicting information was resolved based on the data source (i). In this study, we used information on age, sex, location of detection, exposure history, dates of symptom onset, hospital admission, and official announcement. Details on the collection of individual data and assessment of completeness of variables used in the study are provided in Tab. S1-S2 in Appendix.

We also validated our individual records against the official line lists obtained from the websites of Shandong Provincial Health Commission, Shenzhen Municipal Health Commission, and Hunan Provincial Health Commission for the key variables used in this study (see Tab. S3 in Appendix).

### Statistical analysis

We restricted analyses to the provinces other than Hubei where the majority of our individual records are available (98%, 8,579/8,738). We used the date of one key change in case definition to divide the epidemic into two time periods. The first period runs from the emergence of COVID-19 in Wuhan through to January 27, when the definition of suspected cases in the fourth version issued by China CDC was loosened to capture milder cases (radiographic evidence of pneumonia was no longer necessary, and epidemiological links extended to contact with confirmed cases). The second period runs from January 28 through to February 17. We performed statistical analyses of demographic and epidemiological characteristics of confirmed cases stratified by the two epidemic periods.

We estimated key epidemiologic parameters on time-to-event distributions for COVID-19 cases, including symptom onset to first healthcare consultation, hospital admission, and official announcement. We estimated the time from infection to symptom onset (incubation period) by analyzing COVID-19 cases with asserted epidemiological links (clusters) identified by prospective contact tracing. The date of presumed infection was estimated from cases’ history of exposure, excluding cases with exposure to Wuhan. When multiple exposures were reported, we considered the interval between the first and last recorded dates of exposure. We fitted three parametric distributions (Weibull, gamma, and lognormal) to time-to-event data and selected the best fit based on the minimum Akaike information criterion (AIC).

We analyzed clusters of COVID-19 cases with epidemiological links asserted by prospective contact tracing to estimate the serial interval, which is defined as the interval between onset of symptoms in a primary case and the onset of symptoms in secondary cases generated by that primary case. The serial interval is then estimated by fitting a gamma distribution to the lag between the dates of symptom onset of the primary and secondary cases (with no travel history to/from Wuhan/Hubei) across all clusters.

By leveraging the estimated distribution of the serial interval, we provide estimates of the net reproduction number (*Rt)*, which is the average number of secondary cases generated by a typical primary case at time *t*. Consecutive generations of cases arise after a period measured by the serial interval or by the generation time. We use a Bayesian approach to estimate *Rt* from the time series of symptom onset dates and the distribution of the serial interval^8,9^. For this analysis, the last 9 days of the dataset were not considered to deal with the possible incompleteness of the dataset due to reporting delays. Details on the methodology are reported in the Appendix (page 18).

Statistical analyses were performed with R (version 3.6.0). The reproduction number was estimated from a code in C language written by the authors and available upon request.

### Ethics

The study was approved by the Institutional review board from School of Public Health, Fudan University (IRB#2020-02-0802). All data were collected from publicly available sources and did not contain any personal information.

## Results

As of February 17, 2020, a total of 72,436 COVID-19 cases were reported in all provinces (n=31) of mainland China. 59% (42,752/72,436) of cases including clinically-diagnosed cases (14,953) detected in Wuhan City of Hubei Province, 24% (17,237/72,436) of cases including clinically-diagnosed cases (1,569) in other cities of Hubei Province, and 17% (12,447/72,436) of confirmed cases in other provinces (Fig. 1A). We collected individual information from official publicly available sources on 8,579 laboratory-confirmed cases detected outside Hubei by February 17, accounting for 69% (8,579/12,447) of total cases reported (see Tab.S3 and Fig. S1 in Appendix for a validation of the database and Tab. S4 for an analysis of the representativeness of the dataset). Starting at the end December 2019, the COVID-19 epidemic grew rapidly outside Hubei, characterized by a mix of local transmission and importation of cases from Hubei (Fig. 1B).

**Figure 1.**
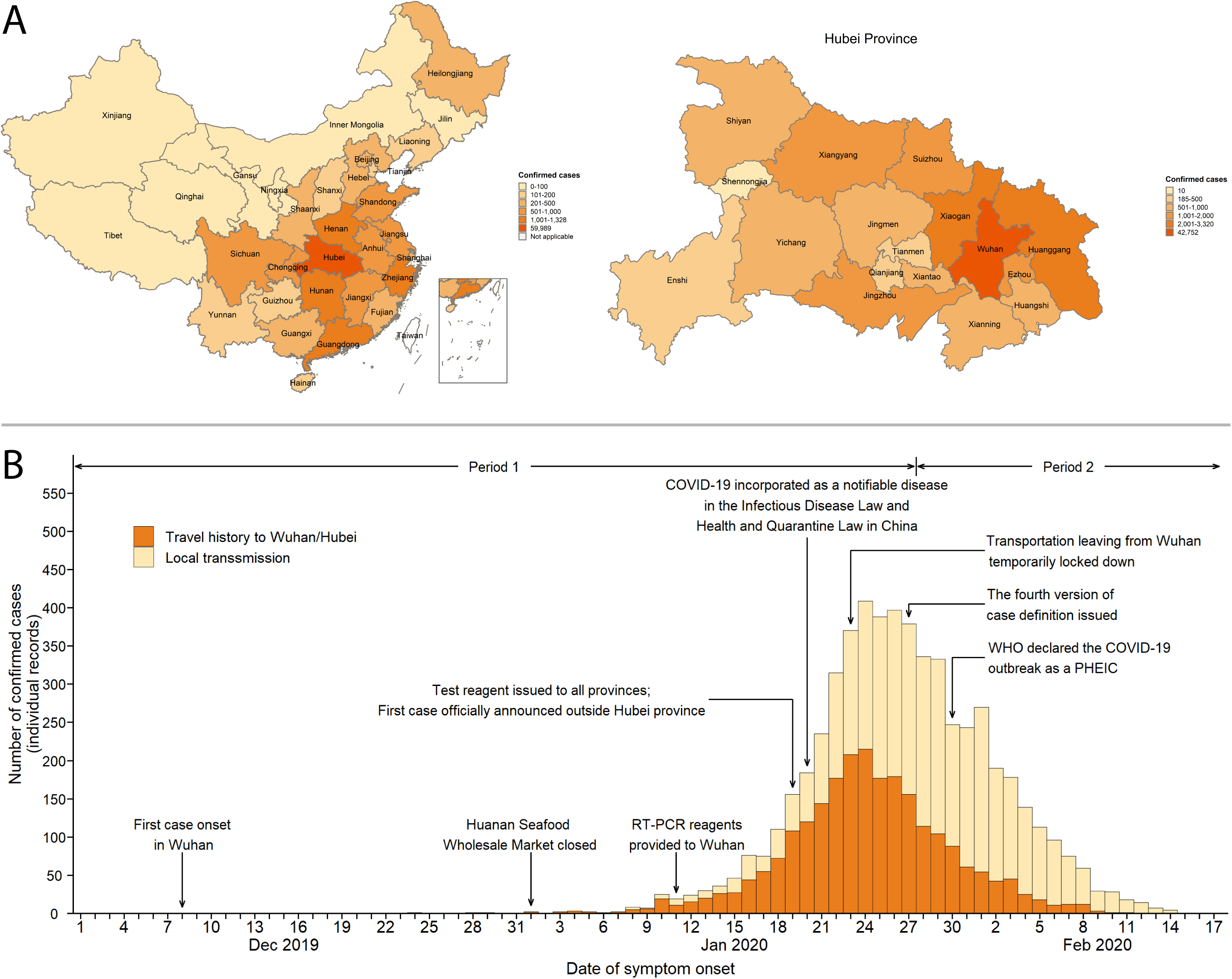
**A** Geographical distribution of confirmed COVID-19 cases (including clinically-diagnosed cases for Hubei Province) officially reported as of February 17, 2020 (N=72,436) in each province of mainland China (left) and in each city of Hubei Province (right). B Time series of COVID-19 cases (date of symptom onset) for which we have individual records in provinces outside Hubei Province, divided into cases with travel history to Wuhan/Hubei and local transmission. Note that if no information about the travel history of a patient is reported in our individual records, we assume that the case locally acquired the infection. Note also that this figure refers to 5,683 cases with non-missing onset date (out of 8,579 individual cases available in our individual records). The decline in the number of cases in the last few days is partially due to the delay between the date of announcement of cases and the date of symptom onset. RT-PCR: reverse-transcription-polymerase-chain-reaction. PHEIC: Public Health Emergency of International Concern.

The median age of the cases was 44 years (range, 1 month to 97 years), with an increasing proportion of cases in age groups below 18 years old (p<0.001) and above 64 years old (p<0.001) observed in the more recent time period. However, the proportion of cases among individuals aged <18 years remains low (5%), see Tab. 1. The proportion of male cases decreased between the two epidemic periods (p<0.001), from 54% to 49%. As of February 17, 51% of cases were male (see Fig. S2 in Appendix).

**Table 1.**
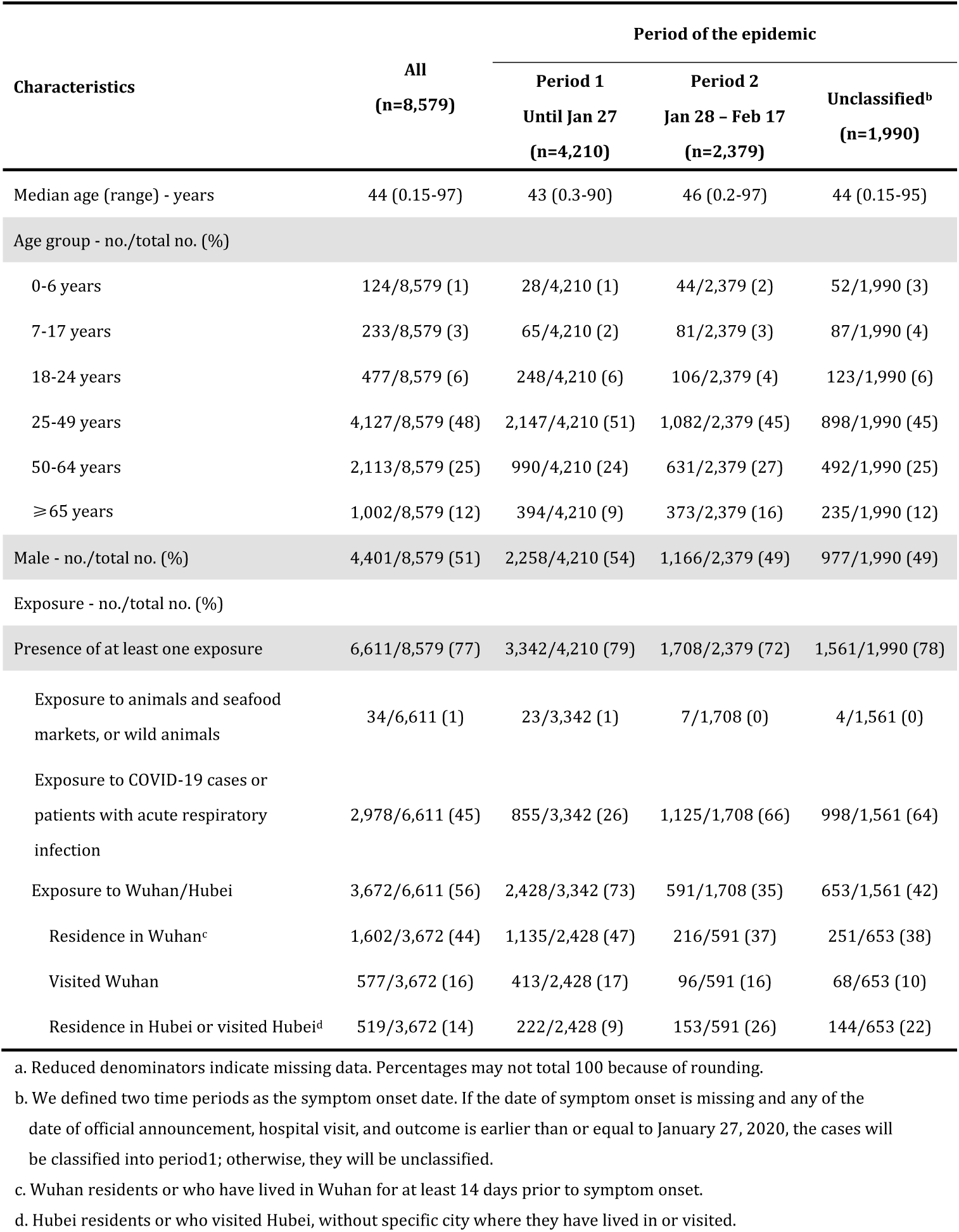
Characteristics of laboratory-confirmed COVID-19 cases in provinces outside Hubei in mainland China by epidemic period, as of February 17, 2020.

| Characteristics | All<br>(n=8,579) | Period of the epidemic |  |  |
| --- | --- | --- | --- | --- |
|  |  | Period 1<br>Until Jan 27<br>(n=4,210) | Period 2<br>Jan 28 – Feb 17<br>(n=2,379) | Unclassified<sup>b</sup><br>(n=1,990) |
| Median age (range) - years | 44 (0.15-97) | 43 (0.3-90) | 46 (0.2-97) | 44 (0.15-95) |
| Age group - no./total no. (%) |  |  |  |  |
| 0-6 years | 124/8,579 (1) | 28/4,210 (1) | 44/2,379 (2) | 52/1,990 (3) |
| 7-17 years | 233/8,579 (3) | 65/4,210 (2) | 81/2,379 (3) | 87/1,990 (4) |
| 18-24 years | 477/8,579 (6) | 248/4,210 (6) | 106/2,379 (4) | 123/1,990 (6) |
| 25-49 years | 4,127/8,579 (48) | 2,147/4,210 (51) | 1,082/2,379 (45) | 898/1,990 (45) |
| 50-64 years | 2,113/8,579 (25) | 990/4,210 (24) | 631/2,379 (27) | 492/1,990 (25) |
| ≥65 years | 1,002/8,579 (12) | 394/4,210 (9) | 373/2,379 (16) | 235/1,990 (12) |
| Male - no./total no. (%) | 4,401/8,579 (51) | 2,258/4,210 (54) | 1,166/2,379 (49) | 977/1,990 (49) |
| Exposure - no./total no. (%) |  |  |  |  |
| Presence of at least one exposure | 6,611/8,579 (77) | 3,342/4,210 (79) | 1,708/2,379 (72) | 1,561/1,990 (78) |
| Exposure to animals and seafood markets, or wild animals | 34/6,611 (1) | 23/3,342 (1) | 7/1,708 (0) | 4/1,561 (0) |
| Exposure to COVID-19 cases or patients with acute respiratory infection | 2,978/6,611 (45) | 855/3,342 (26) | 1,125/1,708 (66) | 998/1,561 (64) |
| Exposure to Wuhan/Hubei | 3,672/6,611 (56) | 2,428/3,342 (73) | 591/1,708 (35) | 653/1,561 (42) |
| Residence in Wuhan<sup>c</sup> | 1,602/3,672 (44) | 1,135/2,428 (47) | 216/591 (37) | 251/653 (38) |
| Visited Wuhan | 577/3,672 (16) | 413/2,428 (17) | 96/591 (16) | 68/653 (10) |
| Residence in Hubei or visited Hubei<sup>d</sup> | 519/3,672 (14) | 222/2,428 (9) | 153/591 (26) | 144/653 (22) |

a. Reduced denominators indicate missing data. Percentages may not total 100 because of rounding.
b. We defined two time periods as the symptom onset date. If the date of symptom onset is missing and any of the date of official announcement, hospital visit, and outcome is earlier than or equal to January 27, 2020, the cases will be classified into period1; otherwise, they will be unclassified.
c. Wuhan residents or who have lived in Wuhan for at least 14 days prior to symptom onset.
d. Hubei residents or who visited Hubei, without specific city where they have lived in or visited.

The presence of at least one known exposure was reported by 77% of cases. In the early epidemic period, most cases reported an exposure in Wuhan or Hubei. Subsequently, an increasing number of cases reported exposure to COVID-19 cases or patients with acute respiratory infections (Tab. 1).

The time interval from symptom onset to hospital admission shortened as the epidemic progressed, decreasing from 4.4 days (95%CI: 0.0-14.0) during the first period of the epidemic to 2.6 days (95%CI: 0.0-9.0) in the second period (Tab. 2). A similar decreasing trend over time was observed for the interval from symptom onset to first healthcare consultation as well (Tab. 2).

**Table 2.** Key time to event intervals of laboratory-confirmed COVID-19 cases by epidemic period, as of February 17, 2020.

| Key time-to-event intervals<sup>a</sup> | All | Period of the epidemic |  |
| --- | --- | --- | --- |
|  |  | Period 1<br>Until Jan 27 | Period 2<br>Jan 28 – Feb 17 |
| Time from symptom onset to first healthcare consultation (days)<sup>a</sup> | 2.5 (95%CI:0.0-10.0),<br>n=2,888 | 3.0 (95%CI:0.0-11.1),<br>n=1,836 | 1.6 (95%CI:0.0-7.0),<br>n=1,052 |
| Time from symptom onset to hospital admission (days)<sup>a</sup> | 3.8 (95%CI:0.0-12.0),<br>n=2,001 | 4.4 (95%CI:0.0-14.0),<br>n=1,310 | 2.6 (95%CI:0.0-9.0),<br>n=691 |
| Time from first healthcare consultation and hospital admission (days)<sup>a</sup> | 1.5 (95%CI:0.0-9.0),<br>n=1,725<sup>b</sup> | 1.4 (95%CI:0.0-9.0),<br>n=850 | 1.4 (95%CI:0.0-9.0),<br>n=353 |
| Time from symptom onset to official announcement (days)<sup>a</sup> | 7.4 (95%CI:1.0-18.0),<br>n=5,024<sup>b</sup> | 8.9 (95%CI:2.0-19.8),<br>n=2,727 | 5.4 (95%CI:1.0-12.0),<br>n=2,079 |

a. Estimated from empirical data; the estimates obtained by fitting gamma, Weibull, and lognormal distributions are reported in Appendix (Tab. S5-S8).
b. Sample size may be different from the sum of the two periods as it includes also cases having not recorded date of symptom onset (which is used to the classification into temporal periods).

We analyzed the time interval from exposure to illness onset for 49 cases (with no travel history to/from Wuhan/Hubei) identified by prospective contact tracing in 37 clusters. We estimated a mean incubation period of 5.2 days (95%CI: 1.8-12.4), with the 95th percentile of the distribution at 10.5 days. We find that the distribution of the incubation period is well approximated by a lognormal distribution (see Fig. S3 and Tab. S9 in Appendix).

We analyzed the time interval between symptom onset in 35 secondary cases and symptom onset in 28 corresponding primary cases. These secondary cases were identified by prospective contact tracing of 28 clusters and had no travel history to Wuhan/Hubei (see Appendix, Fig. S4). One case who reported the onset of symptoms on the same day as the index case was dropped from the analysis (see Appendix, Tab. S10 and Fig. S5 for a sensitivity analysis). Moreover, as we cannot exclude that a fraction of these secondary cases had a previous exposure to an unidentified infection source, we performed a sensitivity analysis by different levels of data censoring (see Appendix, pages 16 and 17). We estimate the serial interval to follow a gamma distribution with a mean of 5.1 days (95%CI: 1.3-11.6). A comparison between the distribution of the incubation period and of the serial interval is reported in Fig. 2.

**Figure 2.**
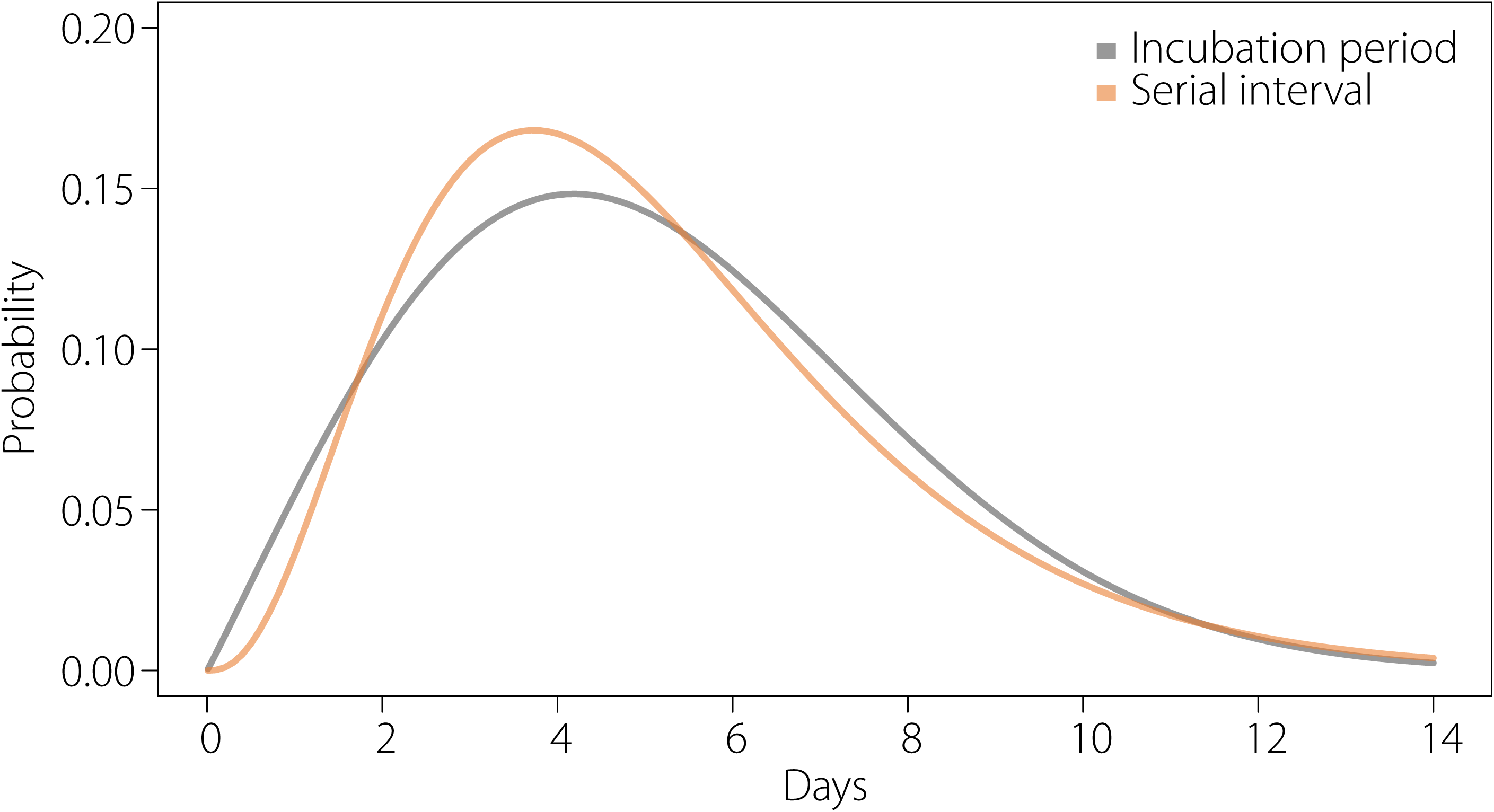
Comparison between the best-fitting distributions of the incubation period and of the serial interval.

The transmission dynamics of COVID-19 are extremely different among the provinces outside Hubei. Here we report the results for a city (Shenzhen – a major city with more than 12 million inhabitants in Guangdong Province) and two Provinces (Hunan and Shandong) for which we have validated our individual records against the full official line list compiled by the respective local health authorities. Results for other seven locations are included in Appendix (Fig. S6 and S7). Despite the selected locations are all among the provinces reporting the largest number of COVID-19 cases as of February 17, 2020^3^, they show highly different transmission patterns. Specifically, in Shenzhen City, we estimated *Rt* above the epidemic threshold for about a week (approximately from January 16 to January 24) with a maximum value of 1.40 (95%CI: 1.04-1.85) when considering a serial interval of 5.9 days on average. The outbreak was mostly sustained by cases with travel history to/from Wuhan/Hubei (Fig. 3A). In Hunan Province, we estimated *Rt* to be above the epidemic threshold for about two weeks with a peak value of 1.58 (95%CI: 1.29-1.92) considering an average serial interval of 5.9 days (Fig. 3B). Shandong Province showed an longer period (more than two weeks) characterized by sustained local transmission and a larger peak value of *Rt*: 2.17 (95%CI: 1.69-2.76) considering an average serial interval of 5.9 days (Fig. 3C). In these three locations, *Rt* remained steadily below the epidemic threshold since the end of January 2020 (Fig. 3A-C). In general, we found that in all the analyzed areas (8 out of 9 most affected provinces outside Hubei and one additional location), *Rt* was below the epidemic threshold as of February 8.

**Figure 3.**
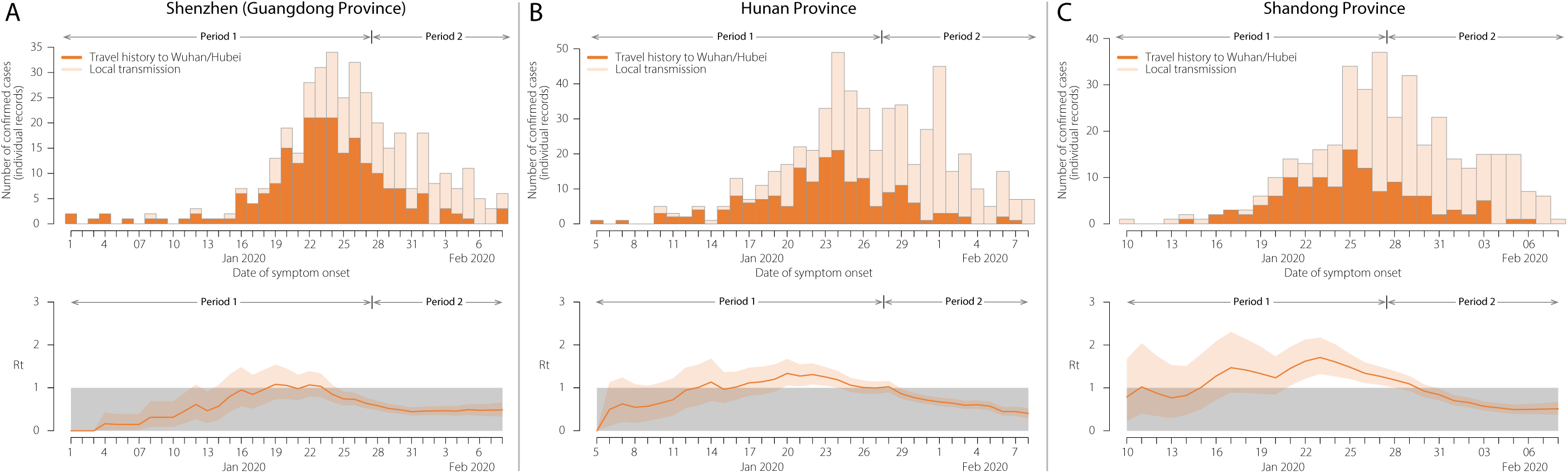
**A** Top: Daily number of new cases (date of symptom onset) in Shenzhen (Guangdong Province) divided into cases with travel history to Wuhan/Hubei and local transmission. Bottom: Estimated net reproduction number (*Rt*) over 4-day moving average. We excluded the last 9 days of data (i.e., data after February 8, 2020) to account for reporting delays. In fact, we estimated the 90^th^ percentile of the distribution of the time from onset to announcement in mainland China outside Hubei Province during period 2 of the epidemic to be 9.0 days. **B** Same as A, but for Hunan Province. **C** Same as A, but for Shandong Province.

## Discussion

We have provided an assessment of the rapidly changing epidemiology and transmission dynamics of the COVID-19 outbreak beyond Hubei Province in mainland China. We found significant differences in the epidemiology of COVID-19 as the epidemic continues to spread across China. Most importantly, as of February 8, 2020, we estimate the net reproduction number to be below the epidemic threshold in the provinces that have reported the largest number of cases (outside Hubei). This suggests that, thanks to the containment measures including isolation of cases, quarantine of contacts, and strict restrictions on personal movement in the general community, China have made key steps towards the interruption of sustained COVID-19 transmission outside Hubei Province.

At the beginning of the epidemic, COVID-19 cases were mostly observed among the elderly^10^. As the epidemic progressed, we observed a shift towards younger (<18 years) and older (65+ years) cases. Since January 28, 2020, however, the proportion of confirmed cases aged <18 years is still only about 5%, although this age group represents approximately 20% of the Chinese population. From the data available here, it is not possible to distinguish whether younger individuals have a reduced risk of infection or a propensity for milder clinical outcome given infection (thus resulting in a lower risk of detection). It should also be considered that schools in China were closed for most of the epidemic due to the 2020 Chinese New Year holidays^11^. It is unclear whether nationwide school breaks could have contributed to the low proportion of confirmed COVID-19 cases among school-age individuals, and whether schools reopening will lead to a change in the transmission patterns of COVID-19.

At the very beginning of the epidemic, a disproportionate faction of COVID-19 cases were male^10^. As of February 17, 2020, however, we estimate that about the same number of cases are observed among males and females (51% of cases are male). This suggests either differential exposure by sex occurring at the beginning of the epidemic (most of cases reported possible exposure to live markets^12,13^) or possible bias in the detection of the first few cases.

In the provinces outside Hubei we estimate the incubation period to be 5.2 days on average, in agreement with previous studies^5,14^. The 95^th^ percentile of the distribution (10.5 days) suggests the importance of contact tracing and medical observations for those with long incubation period.

In the provinces outside Hubei we estimate the serial interval to be on average 5.1 days (95%CI: 1.3-11.6). This estimation is considerably shorter than the preliminary estimate derived from the analysis of six serial intervals in Wuhan^5^. This may be linked to the short time interval from symptom onset to hospital admission (about 2.6 days on average since January 28) we have estimated for mainland China outside Hubei, as compared to what was measured in Wuhan in the early phase of the outbreak^5^, which could have prevented longer serial intervals from being observed. It should however be stressed that, as suggested by a theoretical study^8^, the serial interval estimated from the analysis of household clusters may be up to 20% shorter than the true value.

We estimate the serial interval to have about the same length as the incubation period, in overall agreement with an independent estimate^5^. This suggests the possibility for an early peak of infectiousness with possible transmission before the onset of symptoms. If confirmed, the presence of relevant pre-symptomatic transmission may hamper control efforts, including contact tracing and prompt isolation of index cases, as well as passenger screening at airports. In contrast, strategies based on social distancing and limiting mass gatherings, or contacts in the workplaces and schools, may still be effective^15-17^. Although strong evidence of pre-symptomatic transmission is still lacking, the possibility of transmission during the incubation phase appears to be supported by field epidemiological investigations^18-20^.

The results presented thus far support a change in epidemiological characteristics of the COVID-19 outbreak as time progresses and the epidemic expands to multiple locations. Many of the key epidemiologic time-delay distributions are markedly different from those reported in studies focusing on the early transmission dynamics of COVID-19 in Wuhan^5^. This is likely the consequence of the increased awareness of the public and physicians, behavioral changes of the population with respect to respiratory disease symptoms, increased health care readiness, and elevated alert and response across mainland China.

We estimate the net reproduction number to have followed markedly different patterns in different Chinese provinces. We found that in the analyzed provinces (selected among those mostly affected by the COVID-19 epidemic outside Hubei), the epidemic was self-sustained only for short periods of time (no more than 3 weeks). Most importantly, we estimated that since the end of January 2020, *Rt* is below the epidemic threshold in all the analyzed provinces in mainland China other than Hubei. This is confirmed by the gradual decrease in the number of detected COVID-19 cases reported across all China (outside Hubei Province).

This suggests a beneficial effect of the currently implemented public health intervention policies, in combination with the behavior adopted by the population as a response to the epidemic spread. Nonetheless, it should be stressed that the effectiveness of containment measures only apply while those measures are in place, and a relaxation of public health interventions or a significant change in human behavior may lead to a resurgence in transmission. Moreover, it is important to stress that our findings are based on the analysis of 9 of the most affected provinces (n=10 locations) in mainland China outside Hubei. Therefore, although unlikely, it is possible that in other provinces showing only a few cases at the time of writing the epidemic is still self-sustained by local transmission events. Moreover, it is still unclear to what extent asymptomatic and presymptomatic infections contribute to the transmission of COVID-19 and the level at which they are detected.

It is important to stress that this study is affected by the usual limitations pertaining to the data analysis of rapidly evolving infectious disease outbreaks. The statistical analysis could therefore include biases due to case ascertainment and non-homogenous sampling over time and by location. The level of ascertainment of mild cases remains unclear and could present a different epidemiological characterization. Although the estimates of the net reproduction number are not affected by an approximately constant underreporting rate of cases, they may suffer from time-varying reporting rates. It is also important to consider that the analyzed individual records were retrieved from different data sources and thus may be affected by geographical heterogeneities in sampling of cases with specific exposure (imported or locally acquired infections) and with available dates of symptom onset. However, we analyzed the completeness of individual records used in this study and compared them with official line lists for three locations in mainland China. We found that our data was of similar quality as the official complete line lists.

Even in the presence of the above limitations, a timely updated patient line list like ours is critical to assess the epidemiology and transmission dynamics of an emerging pathogen, inform situational awareness, and optimize the responses to the outbreak. Since January 20, 2020, the National Health Commission of China incorporated COVID-19 as a notifiable disease^21^. The Chinese government committed to timely disclosure of COVID-19 information which was highly praised by the WHO^22^. Accordingly, the local Health Commissions and their officially affiliated medias, where our individual data originates, were authorized to release real-time information about epidemiological investigations on COVID-19 cases. The data collected for this analysis represents a valuable source of information and highlights the importance of publicly available records.

In conclusion, our study provides a detailed overview of the changing epidemiology and transmission dynamics of COVID-19 in mainland China outside Hubei Province. Our findings suggest a slowing down of COVID-19 outbreak in mainland China (outside Hubei Province), indicating that the initial steps taken towards interruption of COVID-19 transmission may be effective. However, the epidemic is not yet under control and a large fraction of the population is still susceptible. The trajectory of the outbreak in China and beyond will depend on the effectiveness of control policies and human behavior in the coming weeks and months.

## Data Availability

All of our data is from public source.

## Contributors

M.A. and H.Y. designed the experiments. J.Z., W.W., Xing.C., X.W., Y.W., X.D., Me.L., W.Z., L.Y., Xinh.C., Q.W., and Y.L. collected data. J.Z., Ma.L., W.W., Y.W., X.D. and M.A. analyzed data. J.Z., Ma.L., W.W., J.Y., K.S., I.M.L., M.E.H., P.W., B.J.C., S.M., C.V., A.V., M.A., and H.Y. interpreted the results. M.A., and H.Y. wrote the manuscript. J.Z., Ma.L., X.W., J.Y., M.E.H., P.W., B.J.C., C.V., and A.V edited the manuscript.

## Declaration of interests

B.J.C. has received honoraria from Roche and Sanofi Pasteur. A.V. has received funding from Metabiota Inc. H.Y. has received research funding from Sanofi Pasteur, GlaxoSmithKline, Yichang HEC Changjiang Pharmaceutical Company, and Shanghai Roche Pharmaceutical Company. None of that research funding is related to COVID-19.

## Acknowledgments

We thank Wenkai Yang, Jingyuan Feng, Jialu Cheng, Qiuyi Xu, Haixin Ju, Xufang Bai, Zi Yu, Yumin Zhang, Wei Guo, Zeyao Zhao, Xin Chen, Sihong Zhao, Rong Du, Jiaxian Chen, Jiangnan Li, Geshu Zhang, Hong Peng, Xin Shen, Zeyu Li, and Yuheng Feng from Fudan University for providing assistance with data collection. We thank Nicole Samay for assistance in preparing the figures.

## Notes

### Funding Statement

H.Y. acknowledges financial support from the National Science Fund for Distinguished Young Scholars (No. 81525023), Key Emergency Project of Shanghai Science and Technology Committee (No. 20411950100), National Science and Technology Major Project of China (No. 2018ZX10201001-010, No. 2018ZX10713001-007, No. 2017ZX10103009-005). M.E.H acknowledges financial support from National Institute of General Medical Sciences U54 GM111274. S.M. and M.A. acknowledge financial support from the European Commission H2020 MOOD project.

## References

1. Chinese Center for Disease Control and Prevention. Epidemic update and risk assessment of 2019 Novel Coronavirus. 2020. http://www.chinacdc.cn/yyrdgz/202001/P020200128523354919292.pdf (accessed Feb 18 2020).

2. National Health Commission of the People’s Republic of China. Update on COVID-19 as of 24:00 on February 17, 2020. 2020. http://www.nhc.gov.cn/xcs/yqtb/202002/261f72a74be14c4db6e1b582133cf4b7.shtml (accessed Feb 18 2020).

3. World Health Organization. Coronavirus disease 2019 (COVID-19) Situation Report – 28 2020. https://www.who.int/docs/default-source/coronaviruse/situation-reports/20200217-sitrep-28-covid-19.pdf?sfvrsn=a19cf2ad_2 (accessed Feb 18 2020).

4. World Health Organization. Statement on the second meeting of the International Health Regulations (2005) Emergency Committee regarding the outbreak of novel coronavirus (2019-nCoV). 2020. https://www.who.int/news-room/detail/30-01-2020-statement-on-the-second-meeting-of-the-international-health-regulations-(2005)-emergency-committee-regarding-the-outbreak-of-novel-coronavirus-(2019-ncov) (accessed Feb 18 2020).

5. Li Q, Guan X, Wu P, et al. Early Transmission Dynamics in Wuhan, China, of Novel Coronavirus–Infected Pneumonia. N Engl J Med 2020.

6. The Novel Coronavirus Pneumonia Emergency Response Epidemiology Team. The Epidemiological Characteristics of an Outbreak of 2019 Novel Coronavirus Diseases (COVID-19) — China, 2020. China CDC Weekly 2020; 2(x).

7. National Health Commission of the People’s Republic of China. Diagnosis and treatment guideline on pneumonia infection with 2019 novel coronavirus (the fifth trial edition). 2020. http://www.nhc.gov.cn/xcs/zhengcwj/202002/d4b895337e19445f8d728fcaf1e3e13a.shtml (accessed Feb 18 2020).

8. Liu Q-H, Ajelli M, Aleta A, Merler S, Moreno Y, Vespignani A. Measurability of the epidemic reproduction number in data-driven contact networks. Proc Natl Acad Sci 2018; 115(50): 12680.

9. World Health Organization Ebola Response Team, Aylward B, Barboza P, et al. Ebola virus disease in West Africa--the first 9 months of the epidemic and forward projections. N Engl J Med 2014; 371(16): 1481–95.

10. Chen N, Zhou M, Dong X, et al. Epidemiological and clinical characteristics of 99 cases of 2019 novel coronavirus pneumonia in Wuhan, China: a descriptive study. Lancet 2020; 395(10223): 507–13.

11. Ministry of Education of the People’s Republic of China. Notice of the Ministry of Education on the postponement of the spring semester of 2020. 2020. http://www.moe.gov.cn/jyb_xwfb/gzdt_gzdt/s5987/202001/t20200127_416672.html (accessed Feb 18 2020).

12. Huang C, Wang Y, Li X, et al. Clinical features of patients infected with 2019 novel coronavirus in Wuhan, China. Lancet 2020; 395(10223): 497–506.

13. Wu P, Hao X, Lau EHY, et al. Real-time tentative assessment of the epidemiological characteristics of novel coronavirus infections in Wuhan, China, as at 22 January 2020. Euro Surveill 2020; 25(3): 2000044.

14. Backer JA, Klinkenberg D, Wallinga J. Incubation period of 2019 novel coronavirus (2019-nCoV) infections among travellers from Wuhan, China, 20-28 January 2020. Euro Surveill 2020; 25(5).

15. Benjamin JC, Eric HYL, Conrad LHL, et al. Effects of School Closures, 2008 Winter Influenza Season, Hong Kong. Emerg Infect Dis 2008; 14(10): 1660.

16. Litvinova M, Liu Q-H, Kulikov ES, Ajelli M. Reactive school closure weakens the network of social interactions and reduces the spread of influenza. Proc Natl Acad Sci 2019; 116(27): 13174.

17. People’s Government of Hubei Province. Notice of the General Office of the People’s Government of Hubei Province on Extending the Spring Festival Holiday of 2020. 2020. https://hubei.gov.cn/zhuanti/2020/gzxxgzbd/zxtb/202002/t20200201_2017564.shtml (accessed Feb 18 2020).

18. Tian HY. 2019-nCoV: new challenges from coronavirus. Chin J Prev Med 2020; 54(0): E001.

19. Yu P, Zhu J, Zhang Z, Han Y, Huang L. A familial cluster of infection associated with the 2019 novel coronavirus indicating potential person-to-person transmission during the incubation period. J Infect Dis 2020.

20. Zou L, Ruan F, Huang M, et al. SARS-CoV-2 Viral Load in Upper Respiratory Specimens of Infected Patients. N Engl J Med 2020.

21. National Health Commission of the People’s Republic of China. Inclusion of 2019 novel coronavirus diseases (COVID-19) into statutory infectious disease management. 2020. http://www.nhc.gov.cn/jkj/s7916/202001/44a3b8245e8049d2837a4f27529cd386.shtml (accessed Feb 18 2020).

22. McNeice A. WHO chief praises containment efforts. 2020. http://www.chinadaily.com.cn/a/202001/31/WS5e3335fba310128217273bff.html (accessed Feb 18 2020).

